# Comparative Analysis of the Duodenojejunal Microbiome with the oral and fecal microbiome unveils its role in Human Severe Obesity

**DOI:** 10.1101/2023.12.15.23299964

**Authors:** Emilie Steinbach, Eugeni Belda, Rohia Alili, Solia Adriouch, Jejunal Luminal and Colonic Mucosa-Associated Microbiota in Metabolic Diseases (Je/Col-MiMe) Group, Benoit Chassaing, Tiphaine Le Roy, Karine Clement

**Author notes:** Corresponding authors. **Correspondence**, Karine Clément, Inserm, Sorbonne University, Nutrition and obesities: systemic approaches (NutriOmics), Sorbonne University, 75013 Paris, France., Tiphaine Le Roy, Inserm, Sorbonne University, Nutrition and obesities: systemic approaches (NutriOmics), Sorbonne University, 75013 Paris, France. these authors contributed equally (Emilie Steinbach, Eugeni Belda).

## Abstract

The intestinal microbiota is recognized as an important player in the development and maintenance of obesity. Most studies focus on faecal microbiota because of its accessibility. However, the small intestine is a major site for nutrient sensing and absorption and few studies have examined the composition and function of the microbiota in this segment of the digestive tract.

We conducted a clinical research project on 30 age- and sex-matched participants with (N=15) and without (N=15) obesity. Duodenojejunal fluid was obtained by aspiration during fibroscopy. Phenotyping included clinical variables related to metabolic status, lifestyle and psychosocial factors using validated questionnaires. Metagenomic analyses of the oral, duodenojejunal and faecal microbiome, as well as metabolomic data from duodenojejunal fluid and faeces, were integrated with clinical and lifestyle data.

The results show associations between duodenojejunal microbiota and lifestyle as well as clinical phenotypes. These associations had larger effect sizes than the associations between these variables and faecal microbiota. We also observed that the duodenojejunal microbiota of obese patients had a higher diversity. In addition, we observed differences in the abundance of several species of the duodenojejunal microbiota between control individuals and patients suffering from obesity.

In conclusion, our results support the relevance of studying the role of the small intestinal microbiota in the development of metabolic diseases.

## Main text

The gut microbiome (GM) plays a pivotal role in the development and maintenance of obesity, however previous research primarily focused on the fecal microbiome (FM). While these studies have provided valuable insights into the compositional and functional changes associated with obesity^1^, the FM represents only a fraction of the gastrointestinal tract (GIT) microbiome. Particularly, the upper small intestine (USI) is of great interest due to its crucial functions in food digestion, nutrient sensing, absorption, enterohormone production, and metabolic homeostasis^2,3^.

Distinct physicochemical conditions exist in each segment of the GIT, shaping microbial ecosystems^4^. Consequently, a comprehensive exploration of the GIT microbiome beyond the FM is warranted. While studies in rodents demonstrated the causal influence of the USI microbiome (USIM) on metabolic regulation, clinical investigations concerning the USIM in human obesity, remain limited and contradictory^3^. There is a pressing need for further research to elucidate the intricate interplay between lifestyle, the USIM, and metabolic health in humans^2,3^.

We investigated the USIM and associated metabolome in thoroughly characterized participants with (n=15, OB) or without (n=15, NOB) obesity matched for age and sex (Table 1). We compared the duodenojejunal fluid (DJF) microbiome aspirated through gastroscopy at the Treitz Angle with the oral microbiome (OM) and FM. Our aim was to uncover the specificities between these microbiomes. Additionally, we conducted statistical analyses to explore potential associations between these microbiomes and participants’ lifestyles and clinical phenotypes.

**Table 1.**
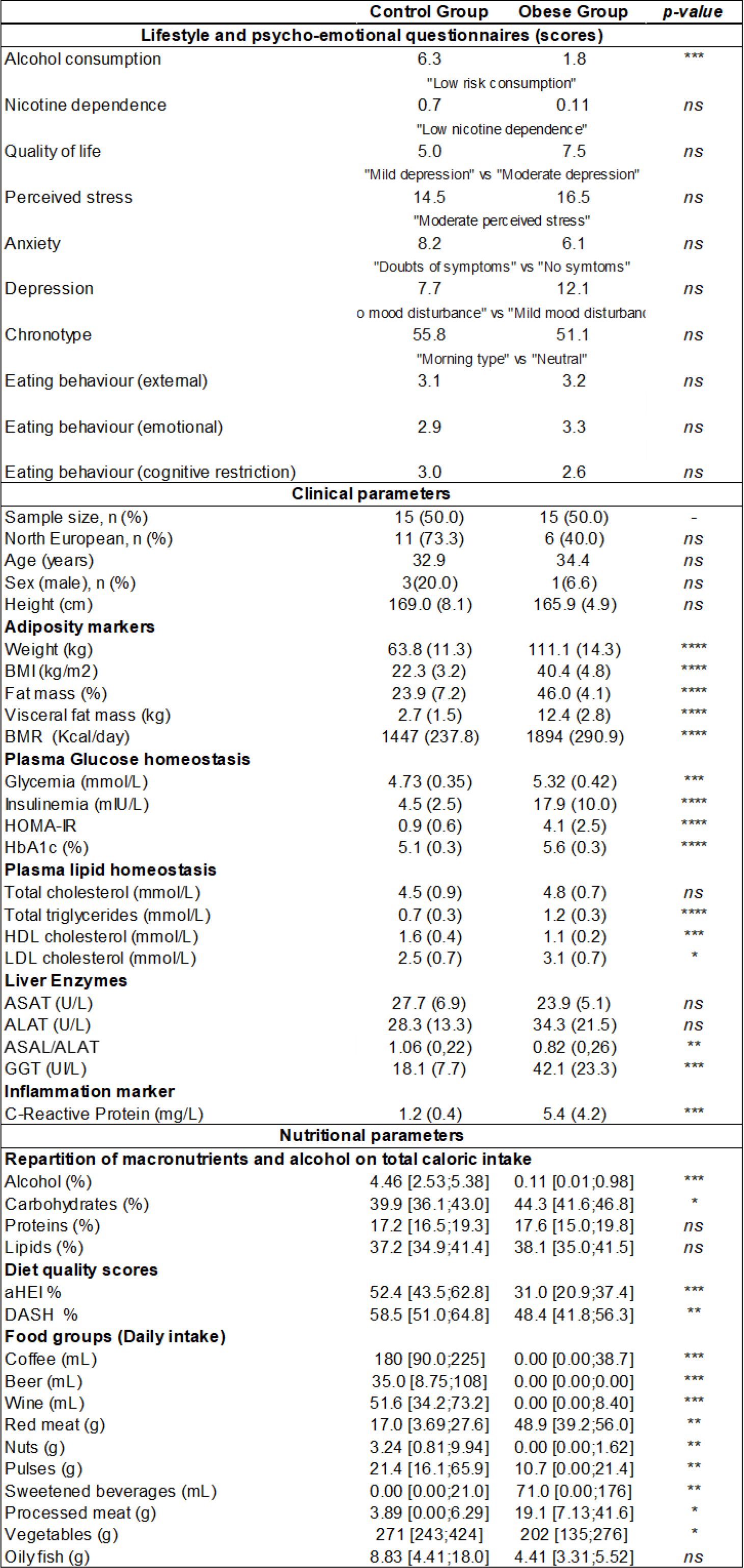
Characteristics of the Study Participants. This descriptive table summarizes the major clinical parameters; the scores obtained from various questionnaires; and some results from the Food Frequency Questionnaire of the Je-MiMe Cohort. Results are expressed as mean (SD) or median [min;max] for continuous data and n (%) for categorical data. Alcohol consumption was evaluated by the Alcohol Use Disorders Identification Test (AUDIT); Nicotine dependence by the Fagerström Test; Quality of Life and Depression by the Patient Health Questionnaire (PHQ-9); Perceived stress by the Perceived Stress Scale (PSS-10); Anxiety by the Hospital Anxiety and Depression Scale (HAD, “A” items only); Depression with The Beck Depression Inventory (BDI); Chronotype or ‘circadian rhythm’ by the Horne and Ostberg Questionnaire; Eating Behavior was assessed with the Dutch Eating Behavior Questionnaire (DEBQ). When available, the corresponding score interpretation is mentioned below the scores. P values result from the Mann-Whitney-Wilcoxon non-parametric test (W test). Legend: BMI: Body Mass Index; HOMA-IR: Homeostatic Model Assessment of Insulin Resistance. ASAT: aspartate transaminase; ALAT: alanine transaminase; GGT: gamma glutamyl-transpeptidase; aHEI (Alternate Healthy Eating Index) and the DASH (Dietary Approaches to Stop Hypertension) scores. ns P-value>0.05; * P-value ≤ 0.05; ** P-value ≤ 0.01; *** P-value ≤ 0.001; **** P-value ≤ 0.0001.

Metagenomic analyses showed that the USIM displayed a lower diversity than the OM and the FM (Figure 1A) and was highly similar to the OM (Figure 1B). This was confirmed by analyzing the distribution of dissimilarities between ecosystems, where dissimilarities between USIM and OM samples was significantly lower than the dissimilarities of both ecosystems vs. fecal samples (Figure 1C). This was further confirmed with a Cliff’s Delta analysis at the species level showing less species being significantly altered between the OM and USIM compared to the FM (Figure 1D). Finally, the prevalence of species altered between the ecosystems showed the presence of aerobic species belonging to the *Streptococcaceae, Veillonellaceae*, and *Prevotellaceae* families in the OM and USIM and the prevalence of strict anaerobes belonging to the *Lachnospiraceae* and *Ruminococcaceae* families in the FM (Figure 1E).

**Figure 1.**
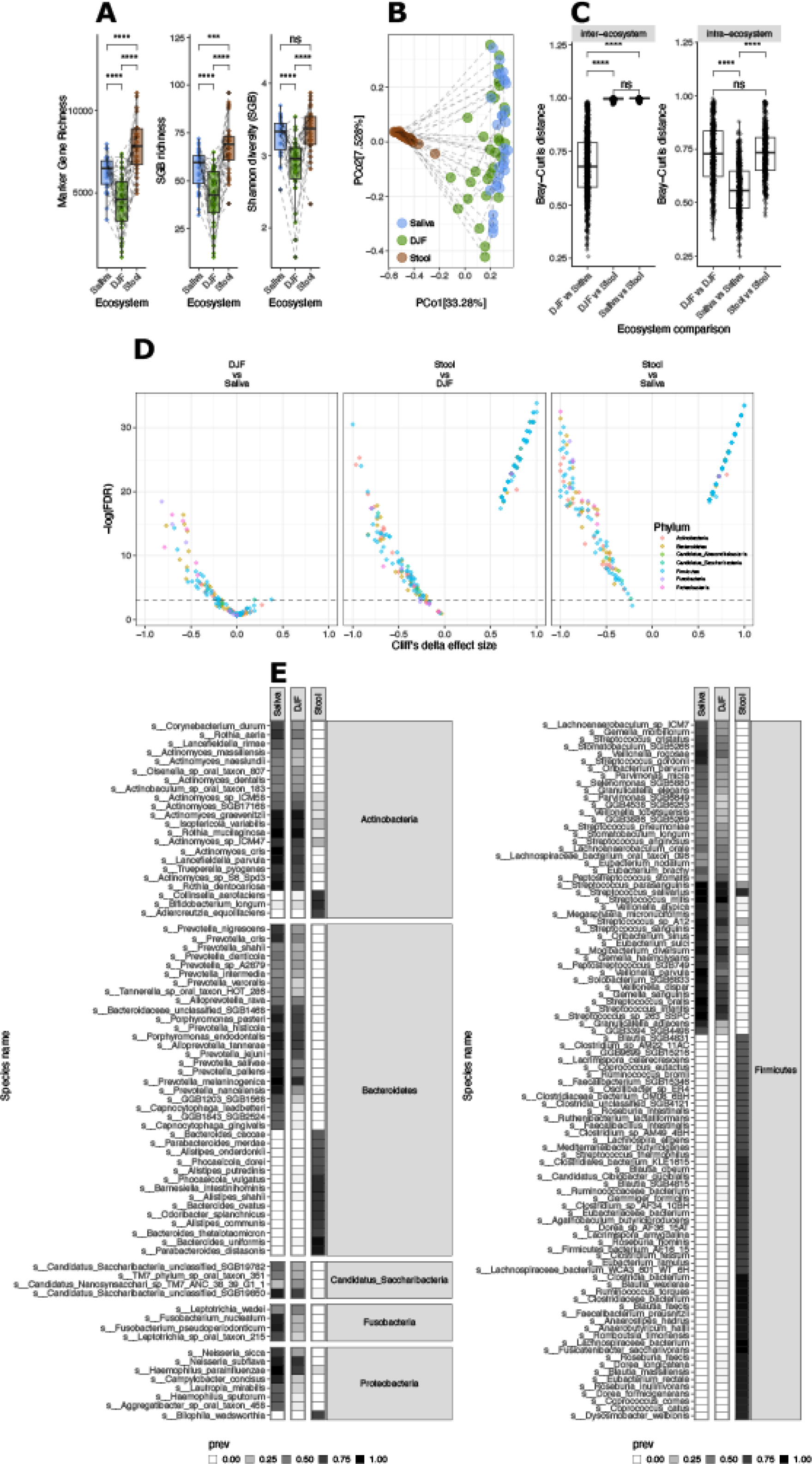
Microbiome patterns across three ecosystems of the digestive tract. (A) <u>Metagenomic richness across the three ecosystems</u>. This boxplot shows the metagenomic richness at the marker gene level (left panel), the species genome bin level (center panel) and Shannon diversity computed at the species genome bin level (right panel; y-axis) of each ecosystem (x-axis): saliva (OM, in blue), duodenojejunal fluid (USIM, in green) and stool (FM, in brown). P-values result from Wilcoxon tests. Legend: **** p<0.0001. (B) <u>Dissimilarity matrix between the three ecosystems</u>. This principal component analysis computed at the species genome bin level shows the distance between each sample (represented by a dot colored in their corresponding ecosystem) in each ecosystem: saliva (in blue), duodenojejunal fluid (in green) and stool (in brown). Samples from the same patient are connected with a dotted line (PCo2). (C) <u>Inter-ecosystem (left panel) and intra-ecosystem (right panel) bray-curtis dissimilarity</u> index computed at the species genome bin level between pairs of ecosystems (left) and within each ecosystem (right). (D) <u>Volcano plot of Cliff’s delta association analysis between species and each ecosystem.</u> Each dot represents a species, colored in their corresponding Phylum. Statistical significance is displayed on the y-axis, and the horizontal dotted line indicates the p < 0.05 threshold. Cliff’s delta is displayed on the x-axis. (E) <u>Species prevalence across the three ecosystems</u>. These heatmaps show the presence-absence (grey-shaded heatmap) of species across ecosystems. Phyla are color-coded using the same legend as panel 1c of this figure. This graph only displays species significantly altered between ecosystems (Kruskall-Wallis: p<0.001). Abbreviations: duodenojejunal fluid (DJF) Fusobacteria (Fuso.) Candidatus Absconditabacteria (C.A.) Candidatus Saccharibacteria (Can. Sacc.).

Comparing within each ecosystem, the groups with or without obesity, we found that the relative abundance *Neisseriaceae* was lower in the USIM of the OB group (Figure 2A). In addition, the USIM in the OB group displayed higher metagenomic richness (Figure 2B) while the other ecosystems richness was similar in NOB and OB group. Coherently, we observed that body composition variables significantly explained the compositional variance of the USIM with a higher effect size than that observed in the OM and FM (Figure 2C) and correlated positively with metagenomic richness (Figure 2D).

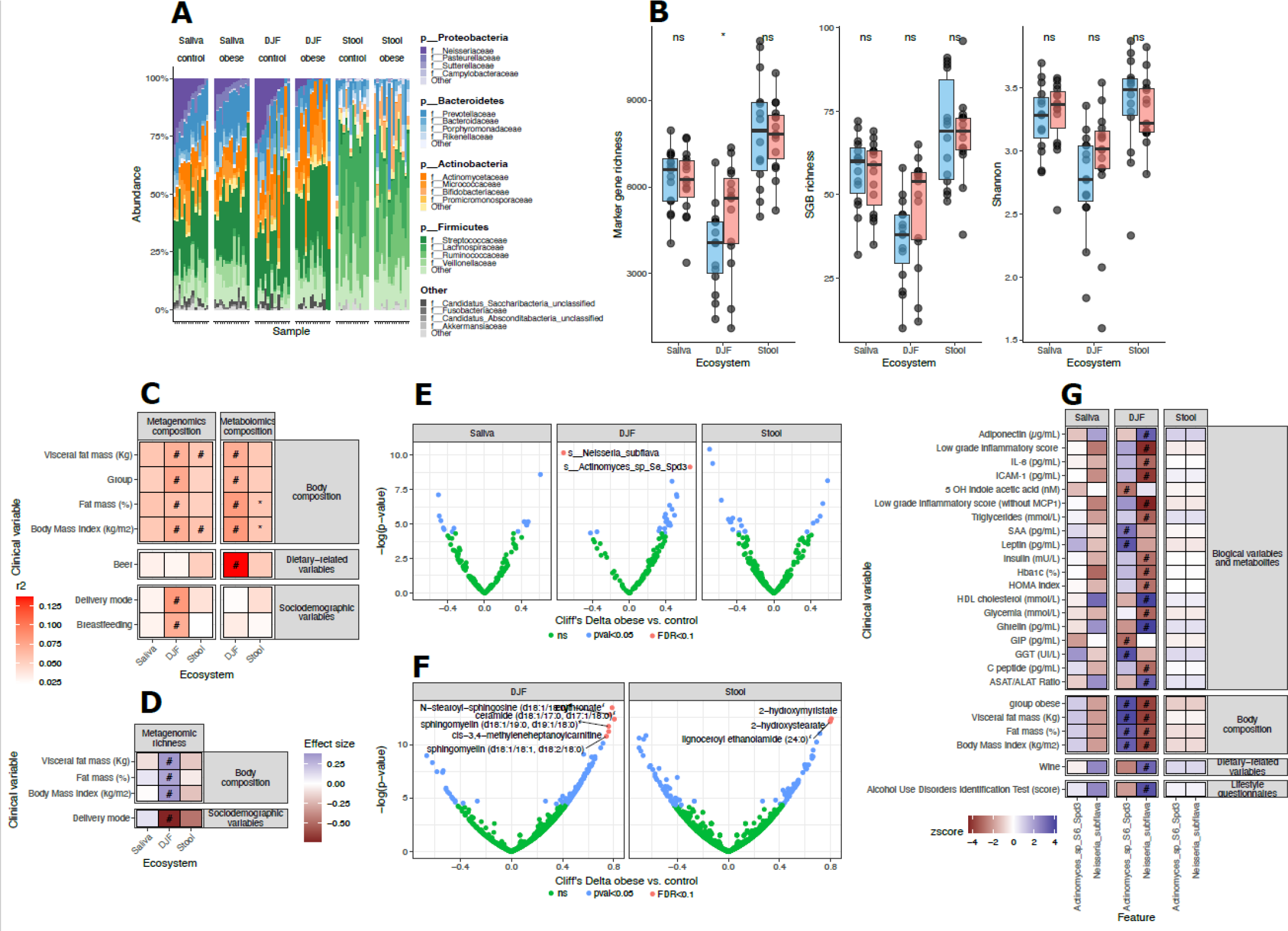
Alterations of the oral, duodenojejunal microbiome and metabolome in obesity and their association with lifestyle and clinical phenotype. (A) Relative abundance (%, y-axis) of bacterial Phyla and families per ecosystem (OM: blue, USIM: green, FM: brown), stratified per group (NOB: group: blue; OB: in red). The different Phyla are listed in bold and color-coded, and bacterial families are colored in different shades within each Phyla. Each column represents a participant. P-values result from between-group comparisons using the Wilcoxon test. (B) Metagenomic richness at the marker gene level (left panel), the species genome bin level (center) and the Shannon index (right panel; y-axis) in each ecosystem (x-axis), stratified per group (NOB group: blue; OB group: red). P-values result from between-group comparisons using the Wilcoxon test. Legend: *p < 0.05. (C) Proportion of compositional variance explained by different clinical or environmental variables. The Permutational Multivariate Analysis of Variance using the Adonis function was computed from the Bray-Curtis dissimilarity matrix for metagenomic data and the Euclidian distances for metabolomics data. The effect size of the determination is color-coded from white (R2 = 0%) to red (R2 = 15%). Legend: #: p < 0.05 and FDR < 0.05. (D) Association analysis between metagenomic richness and lifestyle and clinical variables. This heatmap shows the association between metagenomic richness and lifestyle and clinical variables (listed on the left) in each ecosystem (listed at the bottom) using either Spearman’s rank correlation coefficient, W test or Kruskal–Wallis test. Positive associations are colored in blue, while negative values are colored in red. Legend: Duodenojejunal fluid (DJF)* p < 0.05; # FDR < 0.1. (E) Cliff’s Delta effect-size analysis between bacterial species and participant’s group. P-values are represented on the y-axis, and species’ association to cohort group on the y-axis (NOB: blue; OB: red). Each dot represents a species and is color-coded in regard to the level of significance of the association (green: not significant; blue: p < 0.05; red: FDR < 0.1). (F) Association between metabolites and groups within each ecosystem. These Volcano plots show the Cliff’s Delta effect size analysis between metabolites and the participant’s group within each ecosystem: the DJF (in green) and stool (in brown). P values are represented on the y-axis, and associations with the cohort group on the y-axis (Ctrl in blue, Ob in red). Each dot represents a metabolite and is color-coded in regard to the level of significance of the association (green: not significant - ns; blue: p < 0.05; red: FDR < 0.1). (G) Association analysis between Actinomyces sp S6-Spd3 and Neisseria subflava and clinical and lifestyle variables in the whole cohort of 30 participants (red: negative association; blue: positive association). Legend: # p < 0.05 and FDR < 0.05.

At the species level, 18 species exhibited higher abundance in the OB group USIM, while only three species were more abundant in the NOB group (Figure 2E). Notably, across three ecosystems, 3 species significantly differed between groups after correction for multiple comparisons. In the USIM, *Actinomyces sp* S6-Spd3 was more abundant in participants with obesity, while *Neisseria subflava* was more abundant in the NOB group. In stools, *Ruminococcus lactaris* was more abundant in the NOB group. To determine if USIM alterations in obesity are associated to metabolome changes, we performed a non-targeted metabolome analysis of the DJF and stools, revealing that several lipids and particularly sphingolipids, were increased in the USI of the OB group (Figure 2F). Coherently with the microbiome alterations with obesity within each ecosystem, the USI metabolome variance was more affected by body composition variables than the fecal metabolome (Figure 2C).

Association analyses showed that *Actinomyces sp* S6-Spd3 exhibited positive associations with BMI, fat percentage, visceral fat, along with higher circulating levels of Leptin, Gamma-glutamyl Transferase, and Serum-Amyloid A (Figure 2G). It also displayed a negative association with the circulating levels of gastric-inhibitory-polypeptide. Conversely, *N. subflava* showed negative correlations with corpulence and body composition variables and circulating markers such as glycemia, HbA1c, triglycerides, and inflammatory markers. Additionally, it exhibited positive associations with circulating levels of ghrelin, HDL-cholesterol, adiponectin, and the ASAT/ALAT ratio, as well as lifestyle factors like wine consumption and the Alcohol-Use-Disorder Test score. Further adjusted association analysis highlighted *N. subflava*’s association with BMI even after controlling for alcohol consumption (Figure 2D), suggesting a relationship between *N. subflava* and leanness while considering potential confounding factors.

Our findings support existing studies indicating the similarity between the oral microbiome and the upper small intestine microbiome^5,6^. This contradicts a recent metagenomic analysis across the digestive tract^4^, which reported that the USIM, captured using ingestible capsules, is more akin to the fecal microbiome^4^. These discrepancies may be attributed to several factors, in particular differences in sampling location and variances in collection techniques between studies. Here, we precisely sampled DJF aspirate at the Treitz Angle, while Shalon and colleagues used capsules that may have collected luminal fluid at a less precise and presumably more distal location. In our study, DJF was immediately snap-frozen after sampling, whereas the luminal fluid in the capsules underwent an incubation period until defecation of the capsule. We cannot exclude that the enclosed USIM does not undergo compositional changes during this period.

Previous research has shown reduced diversity in the FM in obesity^7,8^. However, our study indicates the opposite trend in the USIM, suggesting that increased richness is associated with obesity in the upper small intestine. Another report shows an elevated bacterial count in the duodenal mucosa-associated microbiome of hyperglycemic compared to normoglycemic individuals^9^. Further replications in larger cohorts are needed to establish associations between this newly explored microbiome and metabolic health.

Our study, first of its kind, employed whole metagenomic analyses across three digestive tract niches within a meticulously phenotyped cohort, including individuals with obesity and non-obese controls. This work opens avenues for investigating the potential causal impact of two upper small intestinal taxa on metabolic health, particularly in relation to glucose control, inflammation, and body composition.

## Material and methods

### Population

The Je-MiMe study, conducted at Hôpital-Privé des Peupliers, Ramsay-Santé, Paris, France, is an observational study. Prior to inclusion, informed written consent was obtained from all participants. The study adhered to the Helsinki Declaration and ethics committee “Comité de protection des personnes Ile de France VIII” (CPP Ile de France 8; approval number: 210648) of Institut National de la Santé Et de la Recherche Médicale gave approval for this work.

The study comprised 30 participants categorized into two groups: the Non-Obese Group (NOB): composed of individuals without obesity or known metabolic disorders for which gastroscopy was scheduled due to minor epigastralgia that did not necessitate medication; the Obesity Group (OB): composed of candidates for bariatric surgery for which gastroscopy was a prerequisite procedure.

The Je-MiMe study employed specific inclusion and exclusion criteria to select eligible participants and are listed on the Clinical Trial.gov website (NCT05186389).

### Clinical and lifestyle data

Clinical and lifestyle data were collected through online questionnaires. In addition to a general medical questionnaire, various standardized questionnaires were used to evaluate alcohol consumption (Alcohol Use Disorders Identification Test), nicotine dependence (Fagerström Questionnaire), perceived stress (Perceived Stress Scale), anxiety (Hospital Anxiety Depression Scale, only anxiety-items), depression (Beck Depression Inventory), circadian rhythm (Horne and Ostberg Questionnaire), eating behavior (Dutch Eating Behavior Questionnaire), and nutrition (Food Frequency Questionnaire).

Anthropometric measurements and body composition were measured (MC-780MA P, Tanita, Amsterdam, The Netherlands). To avoid redundancy, the results only display a subset of body composition and corpulence variables (fat mass %, visceral fat rating and %, and BMI).

Except for stools, all samples were collected fasting before the gastroscopy, early in the morning. Participants had been fasting for at least 8 hours.

Circulating markers related to glucose metabolism (fasting glycemia, insulin, Hba1c), lipid profile (total cholesterol, LDL, HDL, triglycerides), liver function (aspartate transaminase - ASAT, alanine transaminase - ALAT, gamma-glutamyl transferase and alkaline phosphatase), thyroid function (ultra-sensitive measurement of thyroid-stimulating hormone), and inflammation (C-reactive protein) were measured (Alinity-Abbott; Cerballiance, Paris).

Quantification of circulating levels of Amylin, C-Peptide, Ghrelin, gastric-inhibitory-polypeptide, Glucagon-like Peptide-1, Glucagon, Interleukin-6 (IL-6), Insulin, Leptin, Monocyte-chemoattractant protein-1 (MCP-1), Pancreatic Polypeptide, Peptide-YY, Secretin, and tumor necrosis factor-alpha (TNFα) was performed on serum mixed with dipeptidyl peptidase-4 inhibitors and protease inhibitors (MILLIPLEX® Human Metabolic Hormone Panel V3, Millipore).

Quantification of inflammatory cytokines IL-6 and -8, C-reactive protein, serum-amyloid A, MCP-1, and TNFα were measured on serum (Meso-Scale Discovery’s ultra-sensitive assay). Subsequently, a cumulative score of low-grade inflammation (Zscore) was calculated following the previously described methodology^10^.

Quantitative determination of human High Molecular Weight Adiponectin (Human HMW Adiponectin/Acrp30 Immunoassay), human Growth Differentiation Factor-15, and human Fibroblast growth factor-21 were performed on serum (QuantikineTM, ELISA). Tryptophan metabolites were quantified through liquid chromatography coupled with high-resolution mass spectrometry from serum, as previously described^11^.

### Saliva sampling

Participants were asked not to brush their teeth in the morning before saliva sampling, and they had been fasting for at least 8 hours. After collection, it was transported and aliquoted on ice and stored within two hours at -80°C.

### Duodenojejunal fluid sampling

After saliva sampling, participants thoroughly brushed their mouth and teeth to prevent (as much as possible) DJF contamination from oral cavities with a higher bacterial load^6^. Then, endoscopy was performed. The endoscope was washed in the stomach then DJF was aspirated between the second segment of the duodenum and 10 cm distal to the at the Treitz Angle and collected in a sterile tube. DJF was immediately aliquoted and placed within five minutes after sampling on dry ice, then stored at -80°C.

### Stools sampling

Total fresh stools were collected in a hermetic container at the patient’s home. When the sample was collected, participants placed an anaerocult (bioMérieux, Paris, France) on the stools and hermetically closed the box. The sample was transported and aliquoted on ice in an anaerobic hood within two hours for different analyses, then stored at -80°C.

### Metabolome analysis

Untargeted metabolomics was performed using Ultrahigh Performance Liquid Chromatography-Tandem Mass Spectroscopy on duodenojejunal fluid and stool (Metabolon, Durham, North Carolina, United States).

### Metagenomic analysis

Bacterial DNA extraction from saliva, DJF, and homogenized feces was performed using NucleoMag DNA Microbiome kit (Macherey-Nagel, Vertrieb Gmbh & Co.Kg). Two cycles of chemical- and mechanical-lysis were performed (Precellys®, Bertin Technologies, Montigny-le-Bretonneux, France). We also used an automated robot for DNA extraction and purification using paramagnetic beads (Auto-Pure96, Nucleic Acid Purification System Hangzhou Allsheng Instruments CO., Ltd. Hangzhou, Zhejiang, China). Purity ratio and DNA quantity were controlled (NanoDrop and Qubit, ThermoFisher).

DNA was physically sheared to approximately 250-550 bp through then purified (QIAquick Purification kit, Qiagen, Hilden, Germany). Library preparation for sequencing was performed using the Invitrogen ColibriTM PS DNA Library Prep Kit for IlluminaTM (ThermoFisher Scientific, Waltham, Massachusetts, United States). PCR amplification of the purified adaptor-ligated DNA library was performed, followed by a third purification of the amplified DNA library using reagents included in the Colibri kit. Sequencing was performed with NextSeq 2000 (P2 300 cycles: 2x150 bp).

Metagenomic analyses were performed using the bioBakery tools^12^ for read-level quality control (KneadData, default settings), taxonomic profiling (MetaPhlAn4-catalog^13^ vs mpa_vJan21_CHOCOPhlAnSGB_202103 reference database). To correct for variations in sequencing depth, Metaphlan4 normalized marker gene abundances (reads per kilobase, RPK) were divided by metagenome size (quality-filtered non-human read pairs) before robust average calculation of SGB abundances (0.2 default quantile value).

vegan v2.6.4 R package was used for ecological analyses of metagenomic profiles. Non-parametric statistics (Kruskal-Wallis tests followed by post-hoc pairwise Dunn tests for ecosystem-comparisons; Wilcoxon rank-sum tests for group comparisons) were used to identify taxonomic and metabolomic features associated to different ecosystems and clinical groups. Only features present in >20% of the samples were retained for analyses. P-values derived from KW tests were corrected for multiple testing using the Benjamini–Hochberg method (Padj), only Padj<0.05 were reported as significant. Linear regression analyses were used to evaluate the association of taxonomic and metabolomic markers with clinical covariates unadjusted and adjusted by alcohol intake. All analyses were done on R v4.2.2.

## Supporting information

Descriptive table of the cohort

## Data Availability

All data produced in the present study are available upon reasonable request to the authors. Metagenomics sequencing reads are available on European Nucleotide Archive under Project PRJEB69217.

https://www.ebi.ac.uk/ena/browser/view/PRJEB69217

## Members of the Je/Col-Mime group

Sébastien ANDRE^1^, Emavieve COLES^1^, Laura CREUSOT^6^, Charlène DAURIAT^2^, Gianfranco DONATELLI^3^, Jean-Loup DUMONT^3^, Laurent GENSER^1,4^, Flavien JACQUES^1^, Melissa KORDAHI^2^, Davide MASI^1,5^, Véronique MATEO^1^, Véronique PELLOUX^1^, Christine ROUAULT^1^, Harry SOKOL^6^, Paul TAILLANDIER^1^, Thierry TUSZYNSKI^3^, Marta VAZQUEZ GOMEZ^1^, Jean-Daniel ZUCKER^7^.

^5^ Sapienza University of Rome, Department of Experimental Medicine, Section of Medical Physiopathology, Food Science and Endocrinology, 00161, Rome, Italy.

^6^ Sorbonne Université, INSERM UMRS-938, Centre de Recherche Saint-Antoine, CRSA, AP-HP, Paris, France; Paris Center for Microbiome Medicine, Fédération Hospitalo-Universitaire, Paris, France; INRAE, UMR1319 Micalis & AgroParisTech, Jouy en Josas, France.

^7^ Unité de Modélisation Mathématique et Informatique des Systèmes Complexes, UMMISCO, Sorbonne Universités, Institut de Recherche pour le Développement (IRD), France.

## CRediT Authorship Contributions

Emilie STEINBACH^1^, PhD (Conceptualization: Lead; Data curation: Lead; Formal Analysis: Equal; Funding acquisition: supporting; Investigation: Lead; Methodology: Equal; Project Administration: Lead; Resources - Lead; Software: Supporting; Visualization: Equal; Writing original draft: Lead; Writing – review & editing: Lead).

Eugeni Belda ^2^, PhD (Data curation: Lead; Formal Analysis: Lead; Methodology: Lead; Software: Lead; Validation: Equal; Visualization: Lead; Writing original draft: Supporting; Writing – review & editing: Equal).

Rohia Alili^1^, PhD (Investigation: Equal, Methodology: Equal; Writing – review & editing: Supporting).

Benoit CHASSAING^4^, PhD (Conceptualization: Lead; Investigation: Equal; Methodology: Lead; Project Administration: Equal; Resources: Equal; Supervision: Supporting; Validation: Lead; Writing – review & editing: Supporting).

Tiphaine Le Roy^1^, PhD (Conceptualization: Lead; Investigation: Equal; Methodology: Lead; Resources: Equal; Funding acquisition: Equal; Supervision: Lead; Validation: Lead; Visualization: Equal; Writing original draft: Equal; Writing – review & editing: Lead).

Karine Clément^1,3^, MD, PhD (Conceptualization: Lead; Funding acquisition: Lead; Investigation: Supporting; Methodology: Lead; Project Administration: Equal; Resources: Lead; Supervision: Lead; Validation: Lead; Visualization: Equal; Writing original draft: Equal; Writing – review & editing: Equal).

Solia ADRIOUCH, PhD (Data curation: Equal; Formal Analysis: Equal, Methodology: Equal, Software: Equal; Writing – review & editing: Supporting).

Sébastien ANDRE, PhD, (Investigation: Supporting).

Laura CREUSOT, (Investigation: Supporting).

Charlène DAURIAT (Investigation: Supporting; Methodology: Supporting).

Gianfranco DONATELLI, MD (Conceptualization: Supporting; Investigation: Equal; Project Administration: Supporting; Resources: Equal).

Jean-Loup DUMONT, MD (Investigation: Equal; Resources: Equal).

Laurent GENSER, MD, PhD (Conceptualization: Supporting; Funding acquisition: Supporting; Investigation: Supporting; Project Administration: Supporting; Resources: Supporting).

Flavien JACQUES (Data curation: Equal; Software: Equal).

Melissa KORDAHI, PhD (Investigation: Supporting; Methodology: Supporting; Project Administration: Supporting).

Davide MASI, MD (Investigation: Supporting; Resources: Supporting).

Véronique MATEO, PhD (Investigation: Supporting).

Véronique PELLOUX (Data curation: Equal).

Christine ROUAULT, PhD (Investigation: Supporting).

Harry SOKOL, MD, PhD (Investigation: Supporting; Resources: Supporting).

Paul TAILLANDIER (Investigation: Supporting).

Thierry TUSZYNSKI, MD (Investigation: Equal; Resources: Equal).

Marta VAZQUEZ GOMEZ, PhD (Investigation: Supporting).

Jean-Daniel ZUCKER, PhD (Formal Analysis: Supporting).

## References

1. Aron-Wisnewsky, J., Prifti, E., Belda, E., Ichou, F., Kayser, B.D., Dao, M.C., Verger, E.O., Hedjazi, L., Bouillot, J.-L., Chevallier, J.-M., et al. (2019). Major microbiota dysbiosis in severe obesity: fate after bariatric surgery. Gut 68, 70. 10.1136/gutjnl-2018-316103.

2. Delbaere, K., Roegiers, I., Bron, A., Durif, C., Van de Wiele, T., Blanquet-Diot, S., and Marinelli, L. (2023). The small intestine: dining table of host–microbiota meetings. FEMS Microbiol. Rev. 47, fuad022. 10.1093/femsre/fuad022.

3. Steinbach, E., Masi, D., Ribeiro, A., Serradas, P., Le Roy, T., and Clément, K. Upper small intestine microbiome in obesity and related metabolic disorders: A new field of investigation. Metab. - Clin. Exp. 10.1016/j.metabol.2023.155712.

4. Shalon, D., Culver, R.N., Grembi, J.A., Folz, J., Treit, P.V., Shi, H., Rosenberger, F.A., Dethlefsen, L., Meng, X., Yaffe, E., et al. (2023). Profiling the human intestinal environment under physiological conditions. Nature 617, 581–591. 10.1038/s41586-023-05989-7.

5. Barlow, J.T., Leite, G., Romano, A.E., Sedighi, R., Chang, C., Celly, S., Rezaie, A., Mathur, R., Pimentel, M., and Ismagilov, R.F. (2021). Quantitative sequencing clarifies the role of disruptor taxa, oral microbiota, and strict anaerobes in the human small-intestine microbiome. Microbiome 9, 214. 10.1186/s40168-021-01162-2.

6. Sundin, O.H., Mendoza-Ladd, A., Zeng, M., Diaz-Arévalo, D., Morales, E., Fagan, B.M., Ordoñez, J., Velez, P., Antony, N., and McCallum, R.W. (2017). The human jejunum has an endogenous microbiota that differs from those in the oral cavity and colon. BMC Microbiol. 17, 160. 10.1186/s12866-017-1059-6.

7. Le Chatelier, E., Nielsen, T., Qin, J., Prifti, E., Hildebrand, F., Falony, G., Almeida, M., Arumugam, M., Batto, J.-M., Kennedy, S., et al. (2013). Richness of human gut microbiome correlates with metabolic markers. Nature 500, 541–546. 10.1038/nature12506.

8. Cotillard, A., Kennedy, S.P., Kong, L.C., Prifti, E., Pons, N., Le Chatelier, E., Almeida, M., Quinquis, B., Levenez, F., Galleron, N., et al. (2013). Dietary intervention impact on gut microbial gene richness. Nature 500, 585–588.

9. Darra, A., Singh, V., Jena, A., Popli, P., Nada, R., Gupta, P., Bhadada, S.K., Singh, A.K., Sharma, V., Bhattacharya, A., et al. (2023). Hyperglycemia is associated with duodenal dysbiosis and altered duodenal microenvironment. Sci. Rep. 13, 11038. 10.1038/s41598-023-37720-x.

10. van der Kolk, B.W., Kalafati, M., Adriaens, M., van Greevenbroek, M.M.J., Vogelzangs, N., Saris, W.H.M., Astrup, A., Valsesia, A., Langin, D., van der Kallen, C.J.H., et al. (2019). Subcutaneous Adipose Tissue and Systemic Inflammation Are Associated With Peripheral but Not Hepatic Insulin Resistance in Humans. Diabetes 68, 2247–2258. 10.2337/db19-0560.

11. Lefèvre, A., Mavel, S., Nadal-Desbarats, L., Galineau, L., Attucci, S., Dufour, D., Sokol, H., and Emond, P. (2019). Validation of a global quantitative analysis methodology of tryptophan metabolites in mice using LC-MS. Talanta 195, 593–598. 10.1016/j.talanta.2018.11.094.

12. Beghini, F., McIver, L.J., Blanco-Míguez, A., Dubois, L., Asnicar, F., Maharjan, S., Mailyan, A., Manghi, P., Scholz, M., Thomas, A.M., et al. (2021). Integrating taxonomic, functional, and strain-level profiling of diverse microbial communities with bioBakery 3. eLife 10, e65088. 10.7554/eLife.65088.

13. Blanco-Míguez, A., Beghini, F., Cumbo, F., McIver, L.J., Thompson, K.N., Zolfo, M., Manghi, P., Dubois, L., Huang, K.D., Thomas, A.M., et al. (2023). Extending and improving metagenomic taxonomic profiling with uncharacterized species using MetaPhlAn 4. Nat. Biotechnol. 41, 1633–1644. 10.1038/s41587-023-01688-w.

